# Predictors of healthcare worker burnout during the COVID-19 pandemic

**DOI:** 10.1101/2020.08.26.20182378

**Authors:** Amy V Ferry, Ryan Wereski, Fiona E Strachan, Nicholas L Mills

## Abstract

**Objective:** We aim to provide a ‘snapshot’ of the levels of burnout, anxiety, depression and distress among healthcare workers during the COVID-19 pandemic.

**Design, setting, participants:** We distributed an online survey via social media in June 2020 that was open to any UK healthcare worker. The primary outcome measure was symptoms of burnout as measured using the Copenhagen Burnout Inventory (CBI). Secondary outcomes of depression, anxiety and distress as measured using the Patient Health Questionnaire-9, General Anxiety Scale-7, and Impact of Events Scale-Revised were recorded along with subjective measures of stress. Multivariate logistic regression analysis was performed to identify factors associated with burnout, depression, anxiety and distress.

**Results:** Of 539 persons responding to the survey, 90% were female, and 26% were aged 41-50 years, 53% were nurses. Participants with moderate-to-severe burnout were younger (49% [206/424] *versus* 33% [38/115] under 40 years, P=0.004), and more likely to have pre-existing comorbidities (21% *versus* 12%, P=0.031). They were twice as likely to have been redeployed from their usual role (22% *versus* 11%; adjusted odds ratio [OR] 2.2, 95% confidence interval [CI] 1.5-3.3, P=0.042), or to work in an area dedicated to COVID-19 patients (50% *versus* 32%, adjusted OR 1.6, 95% CI 1.4-1.8, P<0.001), and were almost 4-times more likely to have previous depression (24% *versus* 7%; adjusted OR 3.6, 95% CI 2.2-5.9, P=0.012). A supportive workplace team and male sex protected against burnout reducing the odds by 40% (adjusted OR 0.6, 95% CI 0.5-0.7, P<0.001) and 70% (adjusted OR 0.3, 95% CI 0.2-0.5, P=0.003), respectively.

**Conclusion:** Independent predictors of burnout were younger staff, redeployment to a new working area, working with patients with confirmed COVID-19 infection, and being female or having a previous history of depression. Evaluation of existing psychological support interventions is required with targeted approaches to ensure support is available to those most at risk.

## Introduction

Healthcare workers in both the acute and community settings have played a key role in responding to the global COVID-19 pandemic. As person-to-person transmission was confirmed (1), healthcare workers were faced with increased risk of exposure to SARS-CoV-2 and put under considerable psychological stress with the risk of developing adverse mental health outcomes (2). Previous investigations have shown that direct contact with highly infectious patients is associated with stress in healthcare workers (3-5) therefore burnout among this professional group is of great concern. During the COVID-19 pandemic, healthcare staff have been exposed to increased workload, working in unfamiliar areas, returning to clinical practice from non-frontline roles, pervasive media coverage and concerns about access to appropriate personal protective equipment, layered on top of concerns for the health of family and friends - all factors which could contribute to mental stress. As the wellbeing of health professionals is likely to influence the care they deliver, caring for our staff may also indirectly impact patient outcomes (6).

As the UK along with many countries around the world emerges from lockdown restrictions, now is the ideal time to reflect on the health and wellbeing of our healthcare workforce. Should a wave of cases occur over the winter months and coincide with the annual peak demand on the health service, we will once again call healthcare workers to the front line. Policy makers should be using this opportunity to develop effective support mechanisms to ensure healthcare workers’ mental-wellbeing is prioritised (7).

We aim to provide a ‘snapshot’ of the levels of burnout, anxiety, depression and distress among healthcare workers during the COVID-19 pandemic. Identifying factors associated with increased prevalence of these measures will highlight the need for interventions and identify staff groups who may be more at risk of adverse psychological outcomes. This will better inform future government and health board decisions about allocation of resources to staff wellbeing, and better inform how these resources can be targeted to those with the highest risk of adverse mental-wellbeing outcomes.

## Methods

### Study Population

The study was designed to evaluate the impact of the COVID-19 pandemic on the mental wellbeing of those working in healthcare in the United Kingdom. Anybody working in this sector was eligible for inclusion in the study. Participants were recruited through a non-funded social-media campaign initiated by the study investigators (1 male and 2 female) using snowball sampling (sharing of the survey link among networks). All information was collected anonymously.

### Questionnaire design

The questionnaire collected participant demographics (including age, sex, past medical history, professional role, social living situation and caring responsibilities within the family), concerns over COVID-19 (exposure to patients with COVID-19, access to personal protective equipment, and sources of stress), and access to health and wellbeing support (informal team support, professional support offered by the work place, and external professional support). Questions were developed by the team who were working in clinical roles during this period and reviewed by a clinical psychologist working within the National Health Service (NHS) in Scotland. In addition, validated measures of burnout, depression, anxiety and distress were used to evaluate the incidence of these factors using the Copenhagen Burnout Inventory (CBI) (8), Patient Health Questionnaire-9 (PHQ-9) (9), General Anxiety Disorder-7 (GAD-7) (10) and the Impact of Event Scale (IES-R) (11) respectively. These scales have previously been effective in determining incidence of our chosen outcomes in a healthcare staff population (2, 4, 12, 13). A copy of the questionnaire is available from the authors on request.

### Trial Outcomes

The primary outcome of this study was the incidence of either moderate or severe burnout of healthcare staff. Secondary outcomes included the incidence of either moderate or severe depression, anxiety and distress. All outcomes were based on self-reported symptoms using well-validated measures of burnout, depression, anxiety and distress. A score of 50 or greater on the Copenhagen Burnout Inventory was used as a threshold to indicate moderate or above burnout (calculated as a mean of the three subscales representing total burnout), whereas scores of greater than 10 were used as a threshold for moderate or above depression or anxiety on the Patient Health Questionnaire-9 and General Anxiety Disorder-7 scale. A score above 26 indicated at least moderate distress on the Impact of Events scale. A breakdown for how the scores were calculated is included in the ***Supplementary Appendix* (*Table S1*)**. Additional secondary outcomes included subjective measurement of increased stress during the COVID pandemic, and the reasons for this, as well as the reasons participants may not have accessed support services available in the workplace. Emerging data from Singapore and India suggests that approximately 10% of healthcare professionals managing patients with COVID-19 experience moderate to severe anxiety (14). We estimate that recruitment of 500 participants into our study will identify at least 50 participants who have at least a moderate level of anxiety.

### Ethical Approval

The study was reviewed and approved by the University of Edinburgh ethics review panel from the school of Philosophy, Psychology and Language Sciences.

### Statistical Analysis

Baseline characteristics were summarised for the study population and in groups according to the presence or absence of moderate-severe burnout, depression, anxiety and distress. Continuous variables are presented as mean (SD) or median (IQR), as appropriate. Categorical variables are presented as absolute numbers (%). Group-wise comparisons were performed using Chi-square, Kruskal Wallis or one-way analysis of variance tests as appropriate. Univariable and multivariable logistic regression models were used to estimate the odds of the primary outcome, or either secondary outcome and influence of baseline variables. Baseline variables for the model were selected *a priori* based on their clinical relevance, including age, sex, past medical history (including known ischaemic heart disease or coronary artery disease, diabetes mellitus, or immunocompromise), previous history of mental illness, whether they had a family member at high-risk of COVID-19, whether they were a carer for another adult, as well as descriptors of their working environment (including potential exposure to COVID-19 patients, their access to personal protective equipment, and how supported they felt in their workplace team). All analyses were performed in R (Version 3.5.1).

## Results

### Population demographics

Between 17^th^ and 24^th^ June 2020, 539 participants completed the study questionnaire (26% [141/539] aged 41-50 years, 90% [480/533] female, 53% [286/539] nurses). The majority of participants lived and worked in Scotland (97%, 512/530) ***Table S2***). Accordingly, the occupation and sex of participants in the study closely reflected the National Health Service (NHS) workforce (15). Baseline characteristics of the study population are summarised stratified by the presence or absence of moderate-to-severe burnout (***Table 1***).

**Table 1:** Baseline characteristics of study population stratified by the presence of moderate or severe burnout using the Copenhagen Burnout Inventory. Values as n (%) unless otherwise stated.

|  |  | Moderate-Severe burnout |  |  |
| --- | --- | --- | --- | --- |
|  | All | Present | Absent | P value |
| **No. of participants** | 539 (100) | 424 (79) | 115 (21) | - |
| **Age (years)** |  |  |  |  |
| 21-30 | 121 (22) | 101 (24) | 20 (17) | **0.012** |
| 31-40 | 122 (23) | 104 (25) | 18 (16) |  |
| 41-50 | 141 (26) | 106 (25) | 35 (30) |  |
| 51-60 | 126 (23) | 96 (23) | 30 (26) |  |
| >60 | 27 (5) | 16 (4) | 11 (10) |  |
| **Males** | 53 (10) | 33 (8) | 20 (18) | **0.004** |
| **Significant past medical history** | 104 (19) | 90 (21) | 14 (12) | **0.04** |
| Coronary artery disease | 12 (2) | 10 (2) | 2 (2) | 1 |
| Malignancy | 7 (1) | 6 (1) | 1 (1) | 1 |
| COPD | 60 (11) | 51 (12) | 9 (8) | 0.27 |
| Diabetes mellitus | 16 (3) | 15 (4) | 1 (1) | 0.236 |
| Immunocompromised | 25 (5) | 21 (5) | 4 (4) | 0.677 |
| **Previous mental health diagnosis** | 177 (33) | 159 (38) | 18 (16) | **<0.001** |
| Previous anxiety | 131 (24) | 119 (28) | 12 (10) | **<0.001** |
| Previous depression | 109 (20) | 101 (24) | 8 (7) | **<0.001** |
| Other mental health condition | 21 (4) | 18 (4) | 3 (3) | 0.594 |
| **Family member at high risk** | 99 (19) | 87 (21) | 12 (11) | **0.019** |
| **Main carer for another adult** | 39 (7) | 35 (8) | 4 (4) | 0.125 |
| **Main occupation** |  |  |  |  |
| Allied Health Professional* | 32 (6) | 24 (6) | 8 (7) | **<0.001** |
| Clinical support worker | 54 (10) | 47 (11) | 7 (6) |  |
| Doctor | 57 (11) | 34 (8) | 23 (20) |  |
| Nurse | 286 (53) | 245 (58) | 41 (36) |  |
| Pharmacist | 4 (1) | 2 (1) | 2 (2) |  |
| Other | 106 (20) | 72 (17) | 34 (10) |  |
| **Redeployed from normal workplace** | 105 (20) | 93 (22) | 12 (10) | **0.008** |
| **Exposure to patients with COVID-19** |  |  |  |  |
| No known contact | 94 (17) | 62 (15) | 32 (28) | **<0.001** |
| Patients with suspected COVID-19 | 64 (12) | 43 (10) | 21 (18) |  |
| Patients with confirmed COVID-19 | 131 (24) | 106 (25) | 25 (22) |  |
| Work in a dedicated COVID-19 unit | 250 (46) | 213 (50) | 37 (32) |  |
| **Personal Protective Equipment (PPE)** |  |  |  |  |
| Properly trained with PPE | 300 (57) | 232 (55) | 68 (61) | 0.365 |
| Always have access to PPE when needed | 359 (67) | 273 (64) | 86 (75) | **0.047** |
| Properly fitted for specialist PPE if required | 280 (71) | 225 (71) | 55 (73) | 0.868 |
| Feel safe or very safe when using PPE | 234 (43) | 166 (39) | 68 (59) | **<0.001** |
| **Psychological support** |  |  |  |  |
| Support is available in the workplace | 411 (77) | 308 (73) | 103 (90) | **<0.001** |
| Support was accessed from the workplace if available | 91 (22) | 76 (25) | 15 (15) | 0.064 |
| Support accessed exclusively from outside the workplace | 44 (8) | 36 (9) | 8 (7) | 0.999 |
| Rating of support within team median [IQR]† | 3.0 [2.0, 4.0] | 3.0 [2.0, 4.0] | 4.0 [3.0, 5.0] | **<0.001** |

\* Allied health professional group includes dieticians (n=1), radiographers (n=4), physiotherapists (n=11), and occupational therapists (n=16)
† Rating of support within workplace team is on a self-reported integer scale with 1- least supportive to 5- most supportive.

### Population with moderate-severe burnout

Burnout was present in the majority of participants in the study (79%, 424/539). Compared to participants who did not have moderate-to-severe burnout, those who met the Copenhagen Burnout Inventory criteria tended to be younger (49% [206/424] *versus* 33% [38/115] under 40 years, P=0.004) and female (92% *versus* 83%, P=0.004). They were twice as likely to have pre-existing comorbidities (21% *versus* 12%, unadjusted odds ratio (OR) 1.9, 95% CI 1.4-2.6, P=0.031), and were 4-times more likely to have previous mental illness, such as depression (24% *versus* 7%, unadjusted OR 4.2, 95% CI 2.6-6.1, P<0.001). After adjusting for other variables (***Table 2***), previous depression remained the most powerful independent predictor of moderate-severe burnout, increasing the odds by more than 3-fold (adjusted OR 3.6, 95% CI 2.2-5.9, P=0.012).

**Table 2:** Predictors of moderate-severe burnout. Multivariate logistic regression model adjusted for all variables described with univariate and multivariate odds ratios.

|  | Univariate OR for type moderate-severe burnout (95% CI) | P value for term | Multivariate OR for type moderate-severe burnout (95% CI) | P value for term |
| --- | --- | --- | --- | --- |
| Age |  |  |  |  |
| 20-30 | - | - | - | - |
| 31-40 | 1.1 (0.8-1.6) | 0.703 | 1.2 (0.8-1.8) | 0.64 |
| 41-50 | 0.6 (0.4-0.8) | 0.102 | 0.9 (0.6-1.4) | 0.869 |
| 51-60 | 0.6 (0.5-0.9) | 0.156 | 0.6 (0.4-0.9) | 0.24 |
| 61-70 | 0.3 (0.2-0.5) | 0.007 | 0.4 (0.2-0.8) | 0.174 |
| **Sex (M)** | 0.4 (0.3-0.5) | 0.003 | **0.3 (0.2-0.5)** | **0.003** |
| Significant past medical history*, including immunocompromise | 1.9 (1.4-2.6) | 0.031 | 1.6 (1.1-2.4) | 0.215 |
| Previous anxiety | 3.3 (2.4-4.6) | <0.001 | 1.8 (1.2-2.7) | 0.128 |
| **Previous depression** | 4.2 (2.8-6.1) | <0.001 | **3.6 (2.2-5.9)** | **0.012** |
| Other previous significant mental health condition | 1.7 (0.9-3.1) | 0.426 | 0.9 (0.4-1.9) | 0.856 |
| Family member in a high-risk category of COVID | 2.2 (1.6-3.1) | 0.015 | 1.9 (1.3-2.8) | 0.117 |
| Main carer for another adult | 2.5 (1.4-4.2) | 0.092 | 2.3 (1.1-4.6) | 0.246 |
| **Redeployed from normal working role** | 2.4 (1.7-3.4) | 0.007 | **2.2 (1.5-3.3)** | **0.042** |
| **Exposure level to COVID-19 in working role (per unit increase, scale 1-4)** | 1.5 (1.4-1.6) | <0.001 | **1.6 (1.4-1.8)** | **<0.001** |
| Workplace support available | 0.3 (0.2-0.4) | <0.001 | 0.7 (0.4-1) | 0.314 |
| **Supportive workplace team environment (per unit increase, scale 1-4)** | 0.5 (0.5-0.6) | <0.001 | **0.6 (0.5-0.7)** | **<0.001** |
| Personal Protective Equipment (PPE): |  |  |  |  |
| Adequate training received in use of PPE (Yes) | 0.8 (0.6-1) | 0.311 | 0.8 (0.6-1.1) | 0.482 |
| Access to PPE (scale 1-5) | 0.7 (0.6-0.8) | 0.001 | 0.9 (0.8-1.1) | 0.709 |
| Feeling of safety when using supplied PPE (scale 1-5) | 0.7 (0.6-0.7) | <0.001 | 0.9 (0.8-1) | 0.355 |

\*Significant past medical history includes participants with diabetes mellitus, chronic cardiovascular disease, chronic airways disease, current malignancy, and those receiving oral or intravenous steroids, or immunotherapy agents.

Participant occupation and working environment also had a significant impact on the incidence of burnout (***Table 2***). Those with moderate-severe burnout were twice as likely to have been redeployed from their normal place of work (22% *versus* 11%, adjusted OR 2.2, 95% CI 1.5-3.3, P=0.042), and work in an area dedicated to treating patients with confirmed COVID-19 infection (50% *versus* 32%, adjusted OR 1.6, 95% CI 1.4-1.8, P<0.001). Participants with burnout also had less frequent access to all of the personal protective equipment required to work safely (64% *versus* 75% always having access to personal protective equipment, P=0.047), however in adjusted models this association was attenuated. A supportive workplace team and male sex were both powerful independent predictors protecting against burnout, reducing the odds by 40% (adjusted OR 0.6, 95% CI 0.5-0.7, P<0.001) and 70% (adjusted OR 0.3, 95% CI 0.2-0.5, P=0.003), respectively.

### Secondary outcomes of moderate-severe depression, anxiety, and distress

Baseline characteristics stratified by the presence of moderate-severe depression, anxiety and distress are summarised in ***Table S3***. In participants with moderate or severe depression, as classified by the PHQ-9 score, treating patients with confirmed COVID-19 infection (adjusted OR 1.3, 95% CI 1.1-1.4, P=0.023), previous depression (adjusted OR 3.6, 95% CI 2.6-4.9, P<0.001), and a supportive workplace team (adjusted OR 0.7, 95% CI 0.7-0.8, P=0.001), were all important independent predictors of the odds of depression. Similar results were seen in models to predict moderate-severe anxiety (***Table S4***). Only 5/539 participants scored above the threshold for moderate distress as classified by IES-R (***Table S3***).

### Stress and workplace staff support

The majority of participants (76%, 408/539) recorded that they had felt stressed during the COVID-19 pandemic (***Figure 1***). This was primarily because of concern about their family’s health (73%, 296/408), or their own risk of contracting COVID-19 (64%, 262/408). Other reasons for increased stress included concern about access to personal protective equipment (36% 148/408), their ability to do their job properly (32%, 131/408), and having to cope with higher patient mortality (35%, 144/408), with nearly one third of participants remarking that these reasons had caused them concern. Only a minority of participants who reported stress had accessed local workplace support (23%, 70/306). The most common reason for not doing so was not feeling a need for support, despite increased stress (48%, 113/236), although 38% (89/236) felt they needed support but did not have time, whilst 1 in 7 participants felt that they needed support but they did not want their colleagues to know (14%, 34/236) or that the support was not relevant to them (15%, 36/236). Dedicated workplace support was not available to 1 in 4 participants (25%, 102/408).

**Figure 1:**
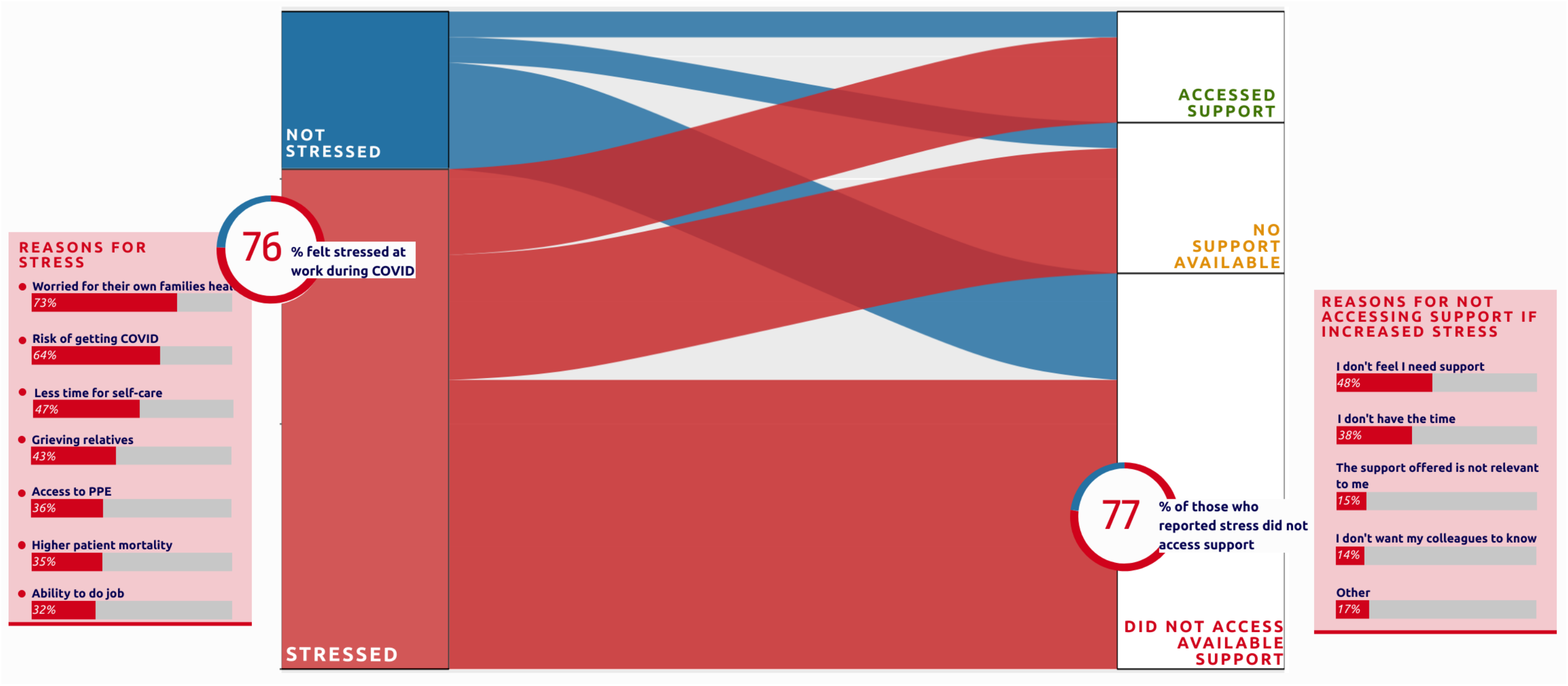
Alluvial diagram showing proportion of participants with increased stress during the COVID-19 pandemic, causes of stress, and whether workplace support was available and accessed. Causes of stress are shown as a stacked bar chart, with size of bar representing proportion, with participants able to select multiple responses.

## Discussion

Prior to COVID-19, burnout among healthcare workers was already a concern (16, 17) The concept of burnout is characterised by feelings of stress due to excessive work demands (exhaustion), a detached attitude towards work and towards those with whom one works (depersonalisation), and reduced feelings of efficiency and attainment (professional efficacy) (18). Working in a healthcare profession during the pandemic may exacerbate these feelings due the unknown nature of the disease, working with a high volume of infected patients, and personal risk of contracting the virus. Burnout not only has consequences of the individual health professional, but also for the patient and colleagues due to higher risk of making poor decisions; possible hostile attitude toward patients; medical errors; and difficult relationships with co-workers (19). Whilst burnout is recognised as a different construct to anxiety and depression, there is an association between the conditions (18). It is imperative that we are aware of these conditions among our workforce for their own personal health, and also their ability to continue to perform their professional roles should a second wave of COVID-19 occur.

We report that the independent predictors of overall burnout among our sample population are being younger, female, redeployed to a new working area, particularly where working with patients with confirmed COVID-19 infection, concerns over access to adequate personal protective equipment, and a prior history of depression.

At the start of a healthcare crisis there is an initial sense of eagerness to contribute to the healthcare effort and a sense of obligation to work through the pandemic (20). As the usual services offered by the NHS were paused to concentrate all available resources on COVID-19, many staff volunteered to work in roles, or were redeployed to roles, that were unfamiliar to them. These redeployed professionals are at particular risk of burnout therefore health and wellbeing interventions should be targeted to reach these staff. Interventions can be focused at the level of the individual or within organisational structures. Organisation directed interventions (teamwork, leadership, workload, shift rotation) were seen to reduce burnout more than individual directed interventions in a meta-analytic review investigating burnout in physicians with De Simone and colleagues concluding that individual directed interventions must be underpinned by organisational approaches (21). Examples of organisational support in a pandemic can take the form of prioritising staff safety (access to personal protective equipment), organising rotas so that teams can stay together, supporting staff to implement recommendations, and ensuring visibility of managers (22). The inadequacy of personal protective equipment supplies was heavily reported in the media at the start of the outbreak and guidance of appropriate personal protective equipment evolved as new knowledge came to light. Keeping the workforce fully informed of the scientific basis for the chosen personal protective equipment provision may help staff feel safer. Psychological interventions targeted at individuals appear to be appreciated by healthcare workers though studies reporting impact on psychological outcomes or experience of using these interventions was rarely reported. Telephone helplines are an example of an intervention targeted at the individual and while staff appreciated the availability of this service, it could also be viewed as too impersonal to be an effective support mechanism (23).

It is interesting to note that more than three quarters of the workforce surveyed reported feeling heightened stress levels at work during the COVID-19 pandemic. Despite this, fewer than 1 in 4 of those with stress had accessed workplace support. The main reason for not accessing support was that they felt they did not require support, despite feeling heightened stress levels.

This phenomenon is not unusual amongst healthcare workers with a delay between event-timing, and reflective practice that allows staff to acknowledge the emotional impact of their work (23). As not all of those recognising their need for support accessed support, this suggests that the structure through which the support is being offered may not be meeting the needs of those it is intended to serve. Not having time to access support, and not wanting colleagues to know they were seeking support were two of the main barriers to access. The use of digital platforms has been shown to be effective in delivering psychological treatment (24). Such platforms may be an option for employers to offer healthcare staff support which can be accessed discreetly at an appropriate time for that individual. The remaining reason for not accessing support were that it was ‘not relevant’. It is possible that the support sought by these individuals related more to organisational structures as described above. These survey results therefore highlight the need for qualitative studies to explore how employers can better support staff from an individual and organisational perspective to identify what type of intervention is desired, what is effective, for whom, and when it is required (23). One of the few factors found to be protective against burnout, depression and anxiety was feeling supported at work. Peer supervision and strong teamwork have been previously shown to strengthen workplace wellbeing (25). Healthcare employers could support service leads, ward managers, senior nurses, and local supervisors to understand their critical role in supporting their staff.

As well as identifying as a healthcare worker, it has been necessary for many staff to also take on a new identity in a way that has not been demanded previously. As patients and families have been unable to be together during critical illness or in the last stages of life, healthcare staff have adopted the role of surrogate family member in very difficult circumstances needing to provide strong emotional support to both patients and relatives. Heavy emotional burden has been previously shown to be a predictor of burnout among healthcare workers (12, 26).

Working in COVID-19 designated areas with a high concentration of critically ill and dying patients will place an even higher emotional demand on staff which may be a contributing cause to burnout seen in this staff group. Focusing support interventions in these areas will be particularly valuable.

As well as a professional identity, healthcare workers also occupy multiple other identities including that of parents, children, siblings, and friends. The major causes of stress among those surveyed were fears for personal health and that of the family. Whilst caring for others, healthcare workers feared putting their own families at risk. Previous studies have reported that prioritising wellbeing of family members represented a barrier to working during an influenza pandemic with degree of worry related to intentional absenteeism (20, 27). With burnout currently present in the majority (79%) of survey participants, employers risk losing staff if not supported adequately should a second wave occur. The COVID-19 pandemic is having a profound impact on society at all levels including mental and physical health (28, 29). Supporting healthcare workers as both individuals and professionals will help optimise individual and team performance in the long term.

Much effort by the public and NHS employers has aimed to help healthcare staff feel supported with offers of gifts or discounts from local and national businesses, a national weekly celebration of their efforts, and relaxation and support hubs in the workplace. As the current health and wellbeing interventions do not appear to be optimised, employers should aim to revise the support offered through thorough evaluation with intended service users and target future interventions to staff groups most at risk. As life in the United Kingdom starts to return to normal, it may not do so for healthcare professionals as they begin to reflect on the impact COVID-19 has had on their professional and personal lives. Taking care of healthcare staff is the only way to ensure their continuing ability to care for our most vulnerable and sick patients.

### Limitations

This study has several limitations. The majority of respondents were female, which is perhaps not unsurprising given than more than half were from the nursing profession where 89% of the workforce are women (15). We did not collect data on ethnicity of respondents. As it is now known that black and ethnic minority groups are more adversely affected by COVID-19 this may have an impact on mental health outcomes among these staff groups. However, in Scotland 96% of the population identify as white (30) therefore it is unlikely we could have performed analyses stratified by ethnicity in our respondent population. Additionally, it is possible that a mental health social media survey has captured the responses of those that are already engaged with the topic and may therefore overestimate the prevalence burnout, anxiety and depression in the target population.

## Data Availability

A de-identified data set will be made available upon request

## Acknowledgments

This study was supported by the British Heart Foundation through a Senior Clinical Research Fellowship (FS/16/14/32023) and a Research Excellence Award (RE/18/5/34216).

